# Sex-Dimorphic Associations of Prior Tuberculosis with Hypertension and Inflammatory Signature in People with HIV: a pilot study

**DOI:** 10.64898/2026.01.12.26343987

**Authors:** Chintu Choolwe Simweene, Frederick sibbenga, Joreen P. Povia, Sepiso K. Masenga

## Abstract

**Background:** People with HIV (PWH) experience an increased burden of cardiovascular disease (CVD), partly driven by chronic immune activation despite antiretroviral therapy (ART). Tuberculosis (TB), a frequent co-infection, may leave persistent inflammatory sequelae even after cure, potentially accelerating hypertension and cardiovascular risk. Evidence on long-term cardiometabolic consequences of prior TB in virally suppressed PWH, particularly sex-specific effects, remains limited.

**Methods:** In a pilot cross-sectional study of 318 PWH, we compared demographics, cardiovascular parameters, and a panel of circulating inflammatory biomarkers between those with and without a history of TB. Logistic regression was used to identify factors associated with TB history in the whole cohort and stratified by sex.

**Results:** In the 31 participants (9.7%) with prior TB, univariate analysis identified significant associations with older age, hypertension, longer ART duration, and elevated IL-6 and soluble ST2. Hypertension was strongly associated with TB history in females (OR 4.41, 95% CI: 3.41 (1.57, 7.41) p=0.003) but not males. In multivariate models adjusted for clinical variables, longer ART duration remained an independent correlate in the full cohort. Sex-stratified multivariate analysis revealed that lower IFN-γ was associated with TB history in males (AOR 0.99, p=0.048), while lower IL-5 was associated with TB history in females (AOR 0.99, p=0.042).

**Conclusion:** A history of TB is associated with hypertension in PWH, particularly among females, and is linked to sex-specific differences in residual inflammatory pathways. These findings suggest that prior TB may contribute to cardiovascular risk in a sex-disparate manner, warranting further investigation.

## Introduction

The global syndemic of human immunodeficiency virus (HIV) and tuberculosis (TB) continues to pose a significant public health challenge, particularly in sub-Saharan Africa (1). While widespread access to antiretroviral therapy (ART) has dramatically reduced AIDS-related mortality, people with HIV (PWH) now face an increased burden of non-communicable diseases (NCDs), including cardiovascular diseases (CVD) (2,3). Chronic immune activation and inflammation, hallmarks of both HIV and treated TB, are believed to be key drivers of this elevated cardiometabolic risk (4,5).

A history of TB disease, even after successful cure, may leave a lasting immunologic scar characterized by persistent low-grade inflammation. This residual inflammatory state, marked by elevated cytokines and cellular activation, can contribute to endothelial dysfunction, arterial stiffening, and the development of hypertension, a major modifiable risk factor for CVD (6). However, the specific inflammatory pathways linking prior TB to hypertension in the context of well-controlled HIV are not well defined. Furthermore, emerging evidence underscores significant biological sex differences in immune responses, inflammation, and cardiovascular pathophysiology (5,7). Women and men may experience distinct trajectories of HIV-associated comorbidity, suggesting that the long-term sequelae of co-infections like TB could also be sex-specific (5).

Despite this, studies investigating the intersection of TB history, chronic inflammation, and hypertension in PWH are scarce, and few have explicitly examined potential sexual dimorphism in these relationships. Most research has focused on active TB or immediate post-treatment outcomes, with less attention paid to the long-term cardiometabolic health of survivors. Addressing this gap is crucial for developing targeted risk stratification and intervention strategies.

Therefore, this pilot study aimed to investigate the hypothesis that a history of TB is associated with a heightened proinflammatory signature and a greater prevalence of hypertension among virally suppressed PWH on stable ART. Secondly, we sought to determine whether the inflammatory and cardiovascular correlates of prior TB differ significantly between males and females. By elucidating these relationships, our findings aim to inform more personalized clinical monitoring and management for PWH with a history of TB.

## Methods

### Study Design and Setting

A pilot cross-sectional study was conducted at the Livingstone University Teaching Hospital Medical Clinic in Zambia from 1^st^ October 2023 1^st^ June 2024. Participants were recruited from the adult HIV clinic during their routine follow-up visits.

### Study Population

The study enrolled adults aged 18 years and older living with HIV who were on stable ART (defined as being on an unchanged regimen for at least 12 months) and were virally suppressed (viral load <1000 copies/mL, or as per national guidelines). Pregnant women and individuals with an active opportunistic infection or acute illness at the time of recruitment were excluded. All participants provided written informed consent.

### Data Collection and Variable Definition

Demographic and clinical data were collected using a standardized questionnaire and from medical records. This included age, sex, body mass index (BMI), waist circumference, documented history of tuberculosis (TB) (defined as a prior diagnosis and completion of anti-TB treatment), duration of known HIV infection and ART, and current blood pressure. Hypertension was defined as a systolic blood pressure ≥140 mmHg and/or diastolic blood pressure ≥90 mmHg on two separate occasions, or a documented diagnosis and current use of antihypertensive medication.

### Biomarker Measurement

Fasting venous blood samples were collected. Plasma levels of inflammatory biomarkers were quantified using an enzyme-linked immunosorbent assay (ELISA). The panel included: high-sensitivity C-reactive protein (Hs-CRP), d-dimer, interferon-gamma (IFN-γ), interleukin (IL)-17A, IL-6, IL-1, IL-5, tumor necrosis factor-alpha (TNF-α), immunoglobulin E (IgE), and soluble ST2.

### Statistical Analysis

Continuous variables were described as medians and interquartile ranges (IQRs) and compared using the Mann-Whitney U test. Categorical variables were described as frequencies and percentages and compared using the Chi-square or Fisher’s exact test. The primary outcome was a history of TB.

Univariate logistic regression was performed to assess the crude association of each variable with a history of TB. Variables with a p-value <0.05 in univariate analysis in the whole cohort were included in an initial multivariate logistic regression model (Model 1). To address Aim 2 on sex differences, all analyses were stratified by sex. A second multivariate model (Model 2) was constructed for each sex, adjusting for clinically relevant variables identified *a priori*: duration of ART and hypertensive status. This approach was chosen to identify inflammatory correlates of TB history independent of these key confounders. All analyses were performed using Statcrunch, a web-based statistical software. A two-sided p-value of <0.05 was considered statistically significant.

### Ethical Considerations

The study was approved by the Mulungushi University School of Medicine and Health Sciences Research Ethics Committee (Reference Number: SMHS-MU3-2023-005, July 9, 2023). All procedures followed were in accordance with the ethical standards of the responsible committee on human experimentation and with the Helsinki Declaration. No data was collected that could potentially identify the participants. All data were de-identified and kept with confidentiality.

## Results

In a cohort of 318 people with HIV (PWH), 31 (9.7%) had a history of tuberculosis (TB). The median age was 49 years (IQR 40-58), and 70.4% were female. Females had a significantly higher BMI and waist circumference than males (p<0.01 for both). There were no significant baseline differences in inflammatory markers or hypertension prevalence by sex (Table 1).

**Table 1.** Demographic and clinical characteristics by Sex.

| Table 1. Demographic and clinical characteristics by Sex |  |  |  |
| --- | --- | --- | --- |
|  | Male | Females | P value |
| Parameter | Median (IQR)<br>OR n(%) | Median (IQR) OR<br>n(%) |  |
| Age (years) | 50 (41, 59) | 48 (40, 58) | 0.362 |
| BMI (kg/m<sup>2</sup>) | 21.9 (19.8, 24.4) | 25.9 (21.8, 29.9) | **<0.0001** |
| Waist circumference (cm) | 78 (72, 88) | 86 (75, 96) | **0.0041** |
| History of TB |  |  |  |
| No | 67 (32.5) | 139 (67.5) | 0.495 |
| Yes | 12 (38.7) | 19 (61.3) |  |
| Hypertension |  |  |  |
| Normotensive | 82 (29.6) | 195 (70.4) | 0.895 |
| Hypertensive | 27 (30.3) | 62 (69.7) |  |
| Duration of HIV/ART (years) | 12.5 (7, 17) | 10 (8, 16) | 0.405 |
| d-dimer (pg/ml) | 3479 (1983, 11417) | 3860 (2068, 13024) | 0.676 |
| Hs CRP (pg/ml) | 67752 (863, 142508) | 75800 (9211, 168744) | 0.344 |
| IFN-g (pg/ml) | 1309 (28, 2213) | 1516 (25, 2346) | 0.609 |
| IL-17A (pg/ml) | 21 (18, 123) | 23 (18, 679) | 0.296 |
| IgE (pg/ml) | 2974 (58, 19614) | 2358 (162, 17108) | 0.945 |
| IL-6 (pg/ml) | 10 (1.0, 108) | 86 (1.0, 1.08) | 0.692 |
| TNF-a (pg/ml) | 1569 (34, 3587) | 577 (27, 3324) | 0.648 |
| IL-1 (pg/ml) | 6.2 (3.1, 183.0) | 3.6 (2.5, 225) | 0.055 |
| IL-5 (pg/ml) | 285 (13, 1114) | 327 (11, 1426) | 0.518 |
| Soluble ST2 (pg/ml) | 3.4 (0.28, 29.20) | **2.6 (0.2, 29.8)** | 0.865 |
| BMI, body mass index; HIV, human immunodeficiency virus; hsCRP, c-reactive protein; IFN-g, interferon gamma; IgE, immunoglobulin E; |  |  |  |

In univariate analysis of the entire cohort, a history of TB was significantly associated with older age (OR 1.07, 95% CI 1.02-1.11, p=0.0007), hypertension (OR 3.41, 95% CI 1.57-7.41, p=0.0019), longer duration of HIV/ART (OR 1.11, 95% CI 1.03-1.19, p=0.003), and higher levels of IL-6 (OR 1.01, 95% CI 1.00-1.02, p=0.027) and soluble ST2 (OR 1.02, 95% CI 1.00-1.05, p=0.003) (Table 2). In a multivariate model adjusting for all significant univariate factors, only longer duration of HIV/ART remained independently associated with a history of TB (AOR 1.18, 95% CI 1.02-1.35, p=0.019). The association with hypertension was attenuated and no longer statistically significant (AOR 3.80, 95% CI 0.96-14.97, p=0.055) (Table 2).

**Table 2.**
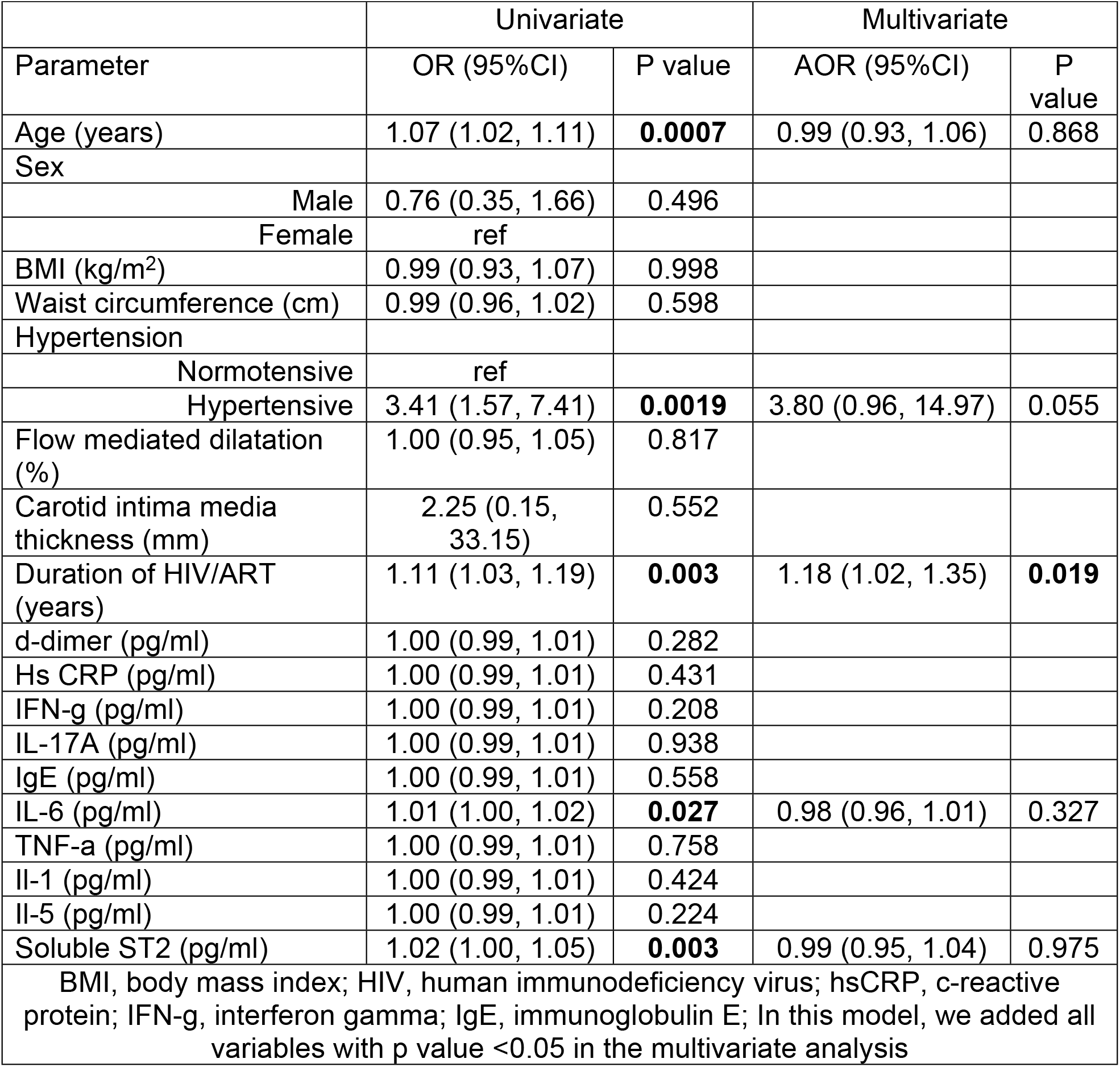
Univariate and multivariate logistic regression of factors associated with history of TB in the whole population.

### Sex differences in correlates of TB history

Sex-stratified univariate analyses revealed distinct patterns (Table 3). Among females, a history of TB was significantly associated with older age (OR 1.06, p=0.007), hypertension (OR 4.41, p=0.003), longer ART duration (OR 1.12, p=0.008), and higher IL-6 (OR 1.02, p=0.0226). Among males, TB history was associated with older age (OR 1.08, p=0.033) and higher soluble ST2 (OR 1.03, p=0.0247), but not with hypertension (OR 2.27, p=0.206). In multivariate models adjusted for clinical variables (Model 2, including ART duration and hypertension), different inflammatory markers were independently associated with TB history by sex: in males, lower IFN-γ was associated with TB history (AOR 0.99, p=0.048), while in females, lower IL-5 was associated with TB history (AOR 0.99, p=0.042) (Table 4).

**Table 3.** Univariate logistic regression of factors associated with TB segregated by sex.

|  |  |  |  |  |
| --- | --- | --- | --- | --- |
|  | Male |  | Female |  |
| Parameter | OR (95%CI) | P value | OR (95%CI) | P value |
| Age (years) | 1.08 (1.00, 1.16) | **0.033** | 1.06 (1.01, 1.12) | **0.007** |
| BMI (kg/m<sup>2</sup>) | 0.97 (-.84, 1.12) | 0.749 | 1.01 (0.93, 1.10) | 0.707 |
| Waist circumference (cm) | 0.98 (0.92, 1.03) | 0.496 | 0.99 (0.96, 1.03) | 0.943 |
| Hypertension |  |  |  |  |
| Normotensive | ref |  | ref |  |
| Hypertensive | 2.27 (0.63, 8.17) | 0.206 | 4.41 (1.63, 11.89) | **0.003** |
| Duration of HIV/ART (years) | 1.08 (0.96, 1.22) | 0.187 | 1.12 (1.03, 1.23) | **0.008** |
| d-dimer (pg/ml) | 1.00 (0.99, 1.00) | 0.437 | 1.00 (0.99, 1.00) | 0.484 |
| Hs CRP(pg/ml) | 0.99 (0.99, 1.00) | 0.203 | 1.00 (0.99, 1.00) | 0.128 |
| IFN-g (pg/ml) | 0.99 (0.99, 1.00) | 0.726 | 1.00 (0.99, 1.00) | 0.247 |
| IL-17A (pg/ml) | 0.99 (0.99, 1.00) | 0.761 | 1.00 (0.99, 1.00) | 0.709 |
| IgE (pg/ml) | 1.00 (0.99, 1.00) | 0.183 | 0.99 (0.99, 1.00) | 0.736 |
| IL-6 (pg/ml) | 1.00 (0.99, 1.01) | 0.597 | 1.02 (1.00, 1.04) | **0.0226** |
| TNF-a (pg/ml) | 0.99 (0.99, 1.00) | 0.335 | 1.00 (0.99, 1.00) | 0.405 |
| Il-1 (pg/ml) | 0.99 (0.99, 1.00) | 0.856 | 1.00 (0.99, 1.00) | 0.134 |
| Il-5 (pg/ml) | 1.00 (0.99, 1.00) | 0.249 | 1.00 (0.99, 1.00) | 0.402 |
| Soluble ST2 (pg/ml) | 1.03 (1.00, 1.07) | **0.0247** | 1.02 (0.99, 1.05) | 0.071 |
| BMI, body mass index; HIV, human immunodeficiency virus; hsCRP, c-reactive protein; IFN-g, interferon gamma; IgE, immunoglobulin E; In this model, we added all variables with p value <0.05 in the multivariate analysis |  |  |  |  |

**Table 4.** Model 2 Multivariate logistic regression of factors associated with TB secreted by sex.

|  |  |  |  |  |
| --- | --- | --- | --- | --- |
|  | MALE |  | FEMALE |  |
| Parameter | AOR (95%CI) | P value | AOR (95%CI) | P value |
| Age (years) | 1.04 (0.96, 1.13) | 0.263 | 1.01 (0.95, 1.08) | 0.593 |
| BMI (kg/m<sup>2</sup>) | 1.01 (0.85, 1.20) | 0.857 | 1.02 (0.93, 1.13) | 0.586 |
| Waist circumference (cm) | 0.98 (0.91, 1.05) | 0.659 | 0.99 (0.94, 1.03) | 0.658 |
| d-dimer (pg/ml) | 1 (0.99, 1.00) | 0.998 | 1.00 (0.99, 1.00) | 0.210 |
| Hs CRP (pg/ml) | 0.99 (0.99, 1.00) | 0.109 | 1.00 (0.99, 1.00) | 0.270 |
| IFN-g (pg/ml) | 0.99 (0.99, 0.99) | **0.048** | 1.00 (0.99, 1.00) | 0.417 |
| IL-17A (pg/ml) | 0.99 (0.99, 1.00) | 0.534 | 0.99 (0.99, 1.00) | 0.722 |
| IgE (pg/ml) | 1.00 (0.00, 1.00) | 0.744 | 0.99 (0.99, 1.00) | 0.403 |
| IL-6 (pg/ml) | 0.93 (0.83, 1.06) | 0.321 | 0.99 (0.96, 1.02) | 0.764 |
| TNF-a (pg/ml) | 0.99 (0.99, 1.00) | 0.238 | 1.00 (0.99, 1.00) | 0.692 |
| Il-1 (pg/ml) | 0.99 (0.99, 1.00) | 0.533 | 1.00 (0.99, 1.00) | 0.554 |
| Il-5 (pg/ml) | 1.00 (0.99, 1.00) | 0.814 | 0.99 (0.99, 0.99) | **0.042** |
| Soluble ST2 (pg/ml) | 1.02 (0.98, 1.06) | 0.187 | 1.01 (0.98, 1.04) | 0.431 |
| In this clinically oriented model 2, in the multivariate analysis, we adjusted all variables for duration on ART, hypertension; BMI, body mass index; HIV, human immunodeficiency virus; hsCRP, c-reactive protein; IFN-g, interferon gamma; IgE, immunoglobulin E; |  |  |  |  |

## Discussion

This pilot study investigated the interplay between a history of TB, inflammation, and hypertension in PWH, with a focus on sex differences. Our primary finding is that a history of TB is associated with markedly increased odds of hypertension in PWH, an association that appears to be driven predominantly by females. Furthermore, we identified distinct sex-specific inflammatory correlates of prior TB infection after adjusting for clinical confounders.

The strong univariate association between TB history and hypertension aligns with growing evidence of the long-term cardiovascular sequelae of TB (8,9). This may be mediated by chronic, low-grade inflammation persisting after cure, which can promote endothelial dysfunction and arterial stiffness. While this relationship was attenuated in the full multivariate model, it remained strong and borderline significant, suggesting it is a clinically important signal warranting investigation in larger cohorts. The independent association of longer ART duration with TB history likely reflects the longer time at risk for acquiring TB prior to or early in the HIV treatment era.

A key strength of this analysis is the sex-stratified approach, which revealed critical differences. The association between TB history and hypertension was significant and substantial in females but not in males. This sexual dimorphism may be influenced by differences in immune responses, body composition (as evidenced by higher BMI and waist circumference in females), or sex-hormone interactions with inflammation and vascular remodeling. The multivariate models further suggest divergent residual inflammatory pathways: prior TB in males was independently associated with lower IFN-γ, a key cytokine for TB immunity, potentially indicating a specific long-term alteration in Th1 response. In females, prior TB was associated with lower IL-5, a cytokine involved in eosinophil activity and humoral immunity, hinting at a different immunologic scar. IL-5 plays a role in eosinophil biology and humoral immune regulation, and its reduction may reflect altered immune homeostasis rather than classical proinflammatory activation. Although IL-5 has been less studied in TB or CVD, emerging evidence suggests that type 2 immune pathways may modulate vascular inflammation differently in women, potentially influencing hypertension risk (5,10). This sex-specific divergence underscores the inadequacy of a one-size-fits-all inflammatory model and highlights the importance of sex-stratified analyses in HIV and TB research.

## Strengths and Limitations

Strengths include a well-characterized cohort, a comprehensive panel of inflammatory biomarkers, and a deliberate analysis of sex differences. A major limitation is the cross-sectional design and limited sample size for participants with a history of TB, which prevents causal inference regarding TB history leading to hypertension or inflammation. Further, the modest sample size, particularly of males with TB history (n=12), limits the power of the sex-stratified multivariate models and increases the risk of Type II error. Reliance on history for TB diagnosis may introduce recall bias, and we cannot ascertain the timing or severity of prior TB infection, which may influence long-term outcomes. Residual confounding from unmeasured factors (e.g., smoking, detailed ART regimens, socioeconomic status) is possible. The results of this study are therefore hypothesis generating providing an avenue for validation.

## Conclusion

A history of TB in PWH is associated with a significantly increased burden of hypertension, a relationship most pronounced in women. Furthermore, the immunologic legacy of prior TB appears to differ by sex, with distinct inflammatory signatures observed in males and females after accounting for key clinical variables. These findings underscore the importance of considering TB history as a potential risk modifier for cardiovascular disease in PWH and highlight the necessity of sex-stratified research. Prospective studies are needed to confirm these associations, elucidate the underlying mechanisms, and determine if targeted screening or interventions for hypertension are warranted in PWH who have survived TB.

## Availability of data and materials

The data are not publicly available due to privacy or ethical restrictions. The data will be made available upon request to qualified researchers, subject to a formal data sharing agreement and approval by our Institutional Review Board, which ensures data will be handled in accordance with the ethical restrictions under which they were collected. To access the data please contact the Mulungushi University School of Medicine and health Sciences Review board, Akapelwa street, Livingstone, Zambia 10101; Phone +260 967758554;

S1 STROBE CHECKLIST

